# Transforming Semi-structured Variant Assessments into Computable Clinical Assertions: A Pilot Study for AI-Assisted Curation

**DOI:** 10.64898/2026.05.07.26352456

**Authors:** Matthew Cannon, Anastasia Bratulin, Kori Kuzma, Daniel Puthawala, Don Corsmeier, Kathleen M. Schieffer, Ben Kelly, Catherine Cottrell, Alex H. Wagner

## Abstract

Genomic medicine relies on expert evaluation of genomic variants, but this process is dramatically slowed by a lack of readily-accessible genomic knowledge. Although genomic knowledge resources such as ClinVar and CIViC support structured data sharing and provide interfaces for adding structure, much of the variant interpretation data generated upstream of these resources is not readily interoperable with these resources, limiting the ability of clinical labs to share data and creating knowledge silos. Here we evaluate a strategy for breaking down these knowledge silos in a pilot study to transform semi-structured variant classification knowledge into computable clinical assertions leveraging the Global Alliance for Genomics and Health (GA4GH) Genomic Knowledge Standards specifications. We programmatically mapped previously captured somatic cancer clinical significance classifications from spreadsheets to the GA4GH Variant Annotation specification. For diagnostic classification data, this approach enabled reuse of standards-aware submission tooling to share 1,499 records to ClinVar. We then studied how AI-assisted curation approaches to overcome gaps in unstructured text enabled scalable curation of prior classifications in unstructured text. Using this approach, we were able to accurately classify clinical significance for 71.8% (117/163) of randomly sampled prognostic evidence statements. We conclude with an overview of how this work may be generalized to make computationally inaccessible variant evidence from other clinical laboratories broadly reusable in downstream knowledgebases such as CIViC and ClinVar.

## BACKGROUND AND SIGNIFICANCE

The adoption of precision oncology has made accurate and scalable genomic variant interpretation a central requirement for effective cancer care. This paradigm depends on the accurate interpretation of individual-specific variants in the context of existing knowledge about their effects across diverse disease settings. An individual who undergoes tumor somatic profiling to characterize their disease may harbor somatic mutations, but without expert interpretation it is often difficult to determine the relevance of any given variant for that individual’s overall molecular profile. To support this effort and provide guidance to clinical directors, pathologists, and variant scientists, the Association for Molecular Pathology (AMP), American Society of Clinical Oncology (ASCO), and College of American Pathologists (CAP) released a joint consensus set of recommendations (the AMP/ASCO/CAP guidelines) to determine the clinical significance of variant-based evidence in cancer or somatic disease^1^. Accordingly, resources such as the National Institute of Health (NIH) ClinVar^2^ platform for human variations in disease and the expert-curated Clinical Interpretation of Variants in Cancer (CIViC) knowledgebase^3^ were developed to capture community knowledge about variants under these guidelines.

Variant evidence interpretation workflows are frequently implemented at the institutional level using a combination of expert guidelines and local standards and practices^4,5^. Consequently, assessment results are captured in heterogeneous formats, including text documents, spreadsheets, user interfaces, and clinical records^6,7^. Even when these data encode similar information, differences in representation and semantics make cross-institutional integration difficult without a unifying standard by which variant classification knowledge is transferred between systems. Unstructured annotations such as these are difficult to systematically transform into structured representations while preserving the original interpretive intent. Even when relevant evidence can be extracted, reconstructing the evaluator’s reasoning, including the clinical significance values^5^ (e.g., whether cited evidence supports or refutes an assertion), remains a significant challenge for FAIR reuse of these data^8^.

Curating variant evidence as structured data is readily achievable but requires specialized software and/or additional curator effort to realize. Manual curation efforts through curation platforms such as those provided by CIViC, ClinGen^9^, and ClinVar support community-crowdsourcing of this problem, but demonstrate the limitations of data volume vs. available curation time. Specifically, while parts of this data capture effort can be automated, transforming the knowledge of a variant classification workflow accurately is time-consuming with limited available human expert effort to handle. This is demonstrated best by the CIViC community evaluation queue, where while 11,300 clinical evidence statements have been added and approved by the community since its opening, as of the time of writing there still remain 6,458 evidence submissions awaiting curation effort^10^. Similarly, of the 821,365 unique multi-submitter variant records in ClinVar (across germline and somatic variant submissions), approximately 20% (162,824 / 658,541) of these are conflicting, lacking the evidence structures needed to procedurally resolve these discrepancies by community experts. And despite substantial contributions from expert curators worldwide, the volume of emerging evidence continues to outpace available curation capacity.^11^ As manual approaches driven by domain experts are the gold-standard and allow for thorough investigation, but cannot keep up with generated data volume, overcoming systemic limitations in the standardization of expert-curated variant classification knowledge is critical towards scalable resolution of discordant interpretation.

Emerging artificial intelligence (AI) technologies offer new opportunities to enhance variant classification workflows and alleviate the significant time demands on expert manual curation. Generative large language models (LLMs) such as ChatGPT or Claude have emerged and demonstrated their ability to both parse and respond to large and complex queries. As LLM use continues to expand in other industries, users and researchers are investigating new ways to integrate generative AI in medical workflows. Recent research has demonstrated the ability of LLMs to pass medical licensing exams without specialized training^12^, and further documents its ability to accurately answer medical diagnostic and therapeutic strategy questions that previously would be considered esoteric to a non-medical professional^13^. These results combined with past work demonstrating LLM potential for multi-step and complex reasoning^14^ suggests further potential for AI’s to be applied towards the assessment of variants for prognostic, therapeutic, and/or diagnostic impact.

Importantly, while these results demonstrate great promise for the use of generative AI in medical workflows, it is imperative to also consider how implementation would affect the patient downstream. The stochastic nature of response generation combined with the complexity of how context can inform medical decision making for any given patient should give pause to workflows that completely remove expert guardrails. Recent clinical expert evaluation of LLM responses to medical questions has highlighted that even when generative answers are “mostly correct”, the incorrect or missing detail still has potential for harm on the patient downstream^15^. Further, LLM responses are often constructed in an authoritative and convincing manner that can obscure inaccuracies or make them difficult to detect without proper medical training. Because medical workflows can directly impact patient care, the consequences of a mistake due to LLM-derived data hallucination (generation of plausible but false information) are too significant to rely solely on AI-driven outputs and thus justify the need for continued expert human engagement.

In this report, we present our pilot of a framework for transforming semi-structured variant assessment data into computable clinical assertions using the Variant Annotation Specification (VA-Spec) of the Global Alliance for Genomics and Health (GA4GH) Genomic Knowledge Standards (GKS) work stream^16,17^. By mapping our semi-structured internal data to these standards, we generated 1,499 structured diagnostic statements and made this publicly available as the largest ever publicly-available collection of pediatric somatic cancer variant interpretations. We then leveraged GKS-compliant community tooling to submit our records to ClinVar, sharing our data publicly and at-scale with the broader variant classification community. Lastly, we demonstrate a pilot AI-assisted curation system to extract and infer clinical significance values for prognostic evidence statements from unstructured text in clinical classification record notes. By utilizing this system, we highlight a pathway for applying modern AI tooling toward the formalization of data previously buried within unstructured free-text. While only an initial-phase study, our work provides a concrete example of how AI tooling can be integrated into existing curation workflows to enhance scalable reuse of expert knowledge.

## RESULTS

### Composition and ingestion of semi-structured variant assessment forms

Variant assessment forms are semi-structured excel sheets used at Nationwide Children’s Hospital (NCH) to support the capture and evaluation of evidence used in variant classification (**Supp. Figure 1**). In addition to case-level individual identifiers, tumor type, genomic coordinates, and associated HGVS expressions, the form contains a list of evidence criteria that are used to classify the variant’s clinical significance based on the tiering system (Tiers I-IV, respectively representing *Strong Clinical Significance*, *Potential Clinical Significance*, *Uncertain Significance*, *Likely Benign*) from the AMP/ASCO/CAP guidelines. The applied criteria captured in these forms allow variant scientists to systematically assess the therapeutic, diagnostic, and/or prognostic impact of a variant for the tumor type under study. Variant level evidence is prioritized, however gene level evidence (e.g. evidence that applies to the overall cancer relevance of the gene, independent of the variation) is also captured and applied. This classification is calculated based on the highest evidence criteria that is met out of Tiers I through IV. Tier I and II evidence is further classified as therapeutic, diagnostic, and/or prognostic clinical significance before ingest (**Supp. Figure 1**). Since individual ClinVar records are structured around these individual types of clinical significance (diagnostic, prognostic, or therapeutic response predictive), a single form could result in a maximum of three separate ClinVar records.

In addition to determining which evidence criteria to apply for an assessment, analysts are instructed to leave comments and supporting evidence for their decision. Directors’ comments are optional; however, when provided, they can override the analysts’ assessment. Although the fields for case information, variant, and tumor type are structured, these fields allow for users to type custom strings, allowing for missing data, errors, and misplaced information. Comments are entirely unstructured and optional, with the only context being the description of the evidence criteria they are associated with.

Variant assessment forms, filled out by clinical variant scientists and approved by a clinical director, were collated and processed utilizing a three-step workflow ending with submission to the ClinVar knowledgebase for public access (**Figure 1**). The input for this workflow included 3,172 forms collected on May 4th, 2026, of which 25 were excluded due to not being in the targeted location (**Figure 2**). We compiled and structured the remaining 3,147 forms into one json file by selecting the relevant sheet, mapping excel labels to structured fields, removing duplicates and empty columns (e.g. due to reanalysis of preexisting cases over time), and normalizing values (uniform time stamps, cleaning whitespace, etc). Individual and sample identifying information are not retained during the record ingestion process.

**Figure 1.**
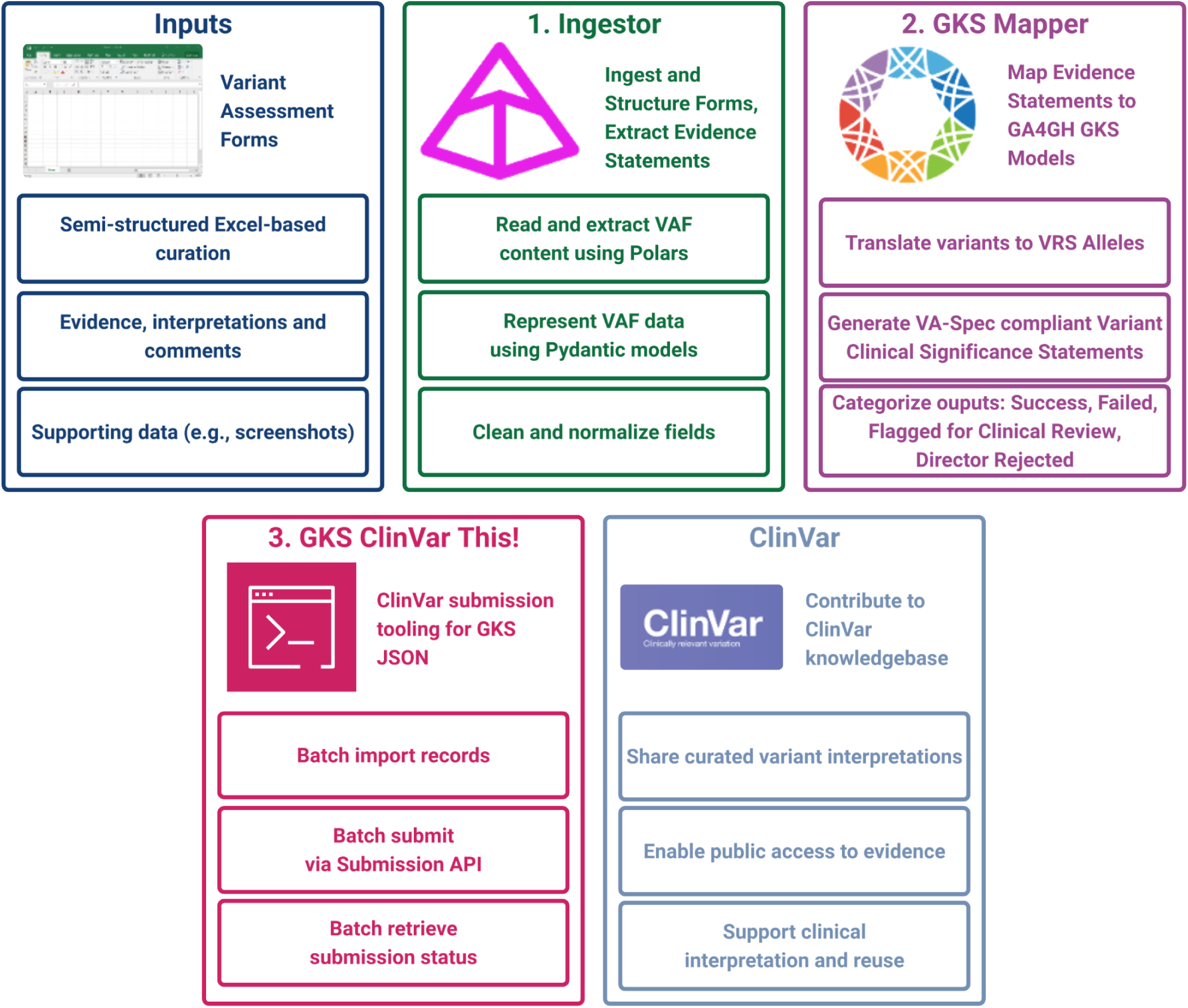
Ingestion of semi-structured variant assessment data for submission to ClinVar. Historical variant assessment data from NCH are available as semi-structured Excel forms containing a mixture of systematic evidence assessments and manually curated evidence. Forms are ingested and structured using Pydantic models, and then are mapped to GA4GH GKS VA-Spec models consistent with the AMP/ASCO/CAP 2017 guideline classifications. Structured GKS representations of variant assessment data are converted to the ClinVar Submission APIschema and submitted via the ClinVar Submission API using *GKS ClinVar This!*.

**Figure 2.**
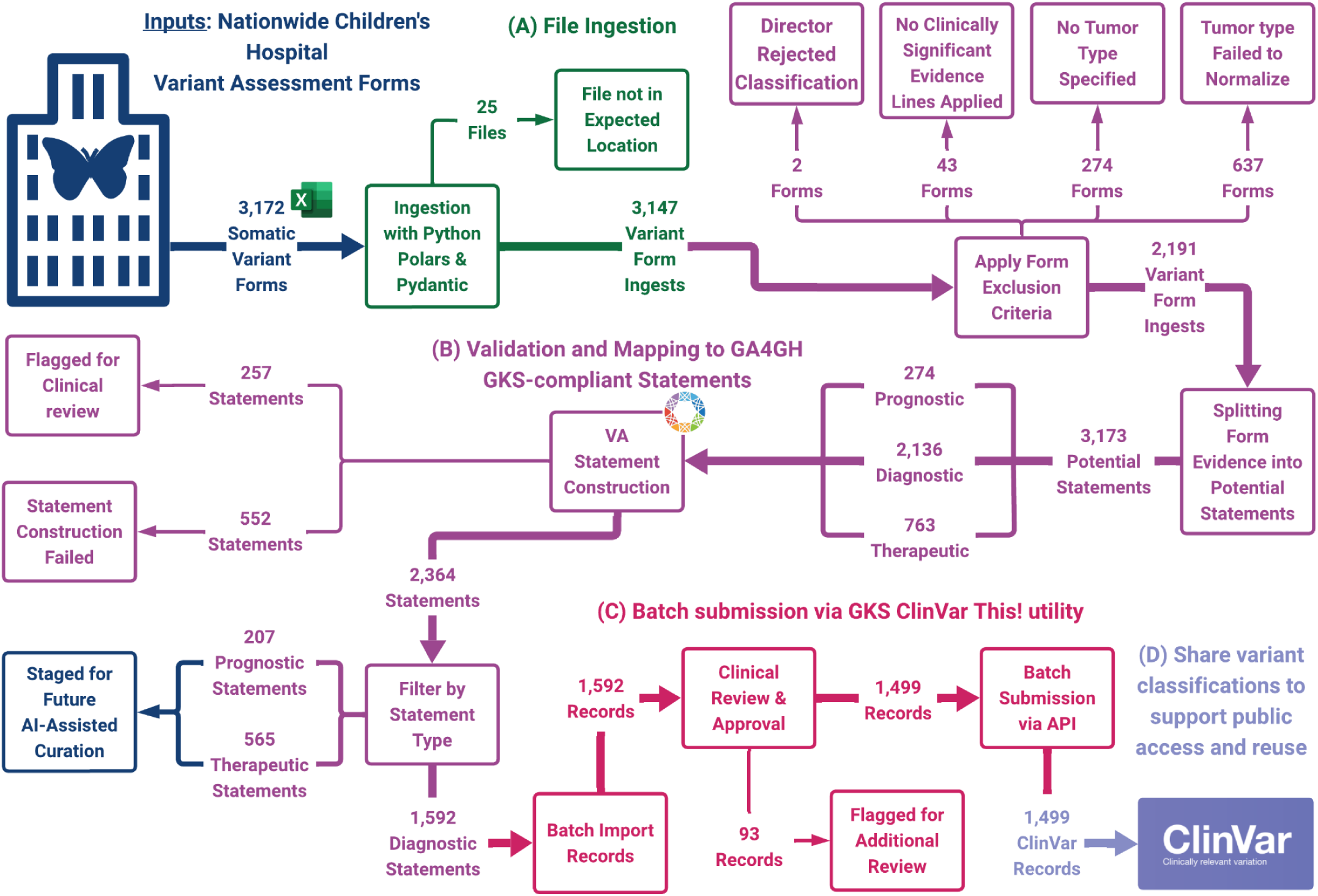
End-to-end workflow for scalable extraction, validation, and submission of clinically relevant variant classifications to ClinVar. (A) Somatic Variant Assessment Forms originating from Nationwide Children’s Hospital serve as the primary input. (B) Files are ingested using a Python-based pipeline (utilizing Polars and Pydantic) to ingest structured and semi-structured content. (C) Evidence line validation filters out records lacking essential components (e.g., variant or tumor type), with invalid or incomplete entries flagged for review. (D) Clinically relevant statements are extracted from validated evidence lines, including diagnostic assertions and deduplicated by variant–tumor type pairs. (E) Extracted diagnostic evidence statements are mapped to GA4GH Genomic Knowledge Standards (GKS), including normalization to VRS Alleles and construction of VA-Spec–compliant diagnostic statements. Failures at this stage are flagged for additional review or rejection. (F) Structured records are submitted via the ClinVar “GKS ClinVar This!” tool, including batch import, clinical review and approval, and API-based submission. (G) Submitted records appear in ClinVar to support public access, reuse, and downstream integration of these variant classifications.

### Filtering and mapping of Variant Assessment Data

The 3,147 ingested forms were filtered for those meeting the minimum data requirements for GA4GH GKS Variation Annotation Specification (VA-Spec) statements and ClinVar clinical impact records. The GKS mapper workflow normalizes variant, tumor type data, and variant relationships, and flags idiosyncratic records for manual review. After normalization, variant and disease concepts were mapped to standardized forms using the Variation Representation Specification (VRS) Python package and the Variant Interpretation for Cancer Consortium (VICC)^18^ Disease Normalizer. A curated internal disease mapping was created by the clinical team as the initial normalization method, followed by the Disease Normalizer (**Supp. Figure 2**).

At this stage, 956 forms were excluded due to clinical director rejection or insufficient data for constructing VA-Spec evidence statements. These exclusions included 2 director-rejected forms, 43 forms lacking clinically significant evidence, 274 forms without a defined tumor type, and 637 forms with tumor types that failed normalization. Those without a tumor type primarily reflected earlier versions of the form that did not include this field. Across the dataset, 629 unique tumor types were observed, of which 296 were successfully normalized.

Statements without defined variants included 281 representing generic loss-of-function assertions, 4 gain-of-function, 2 internal tandem duplications, and 40 with unspecified variant type.

From the remaining 2,191 ingests, we derived 3,173 potential evidence statements, including 2,136 diagnostic, 763 therapeutic, and 274 prognostic statements. During VA-Spec statement construction, 552 statements failed, including 327 lacking a defined variant, 77 with variants that could not be mapped to VRS alleles, and 148 that were not VA-Spec compliant. Statements without defined variants included 281 representing generic loss-of-function assertions, 4 gain-of-function, 2 internal tandem duplications, and 40 with unspecified variant type.

An additional 126 statements were collapsed as duplicate variant-tumor type pairs, retaining the most recent instance. These retained statements contributed to a set of 131 statements flagged for review, which also included cases with disease mapping uncertainty or variant normalization issues.

In total, 2,364 statements were successfully constructed, of which 1,592 diagnostic statements were eligible for submission via the *GKS ClinVar This!* tool. The remaining 565 therapeutic and 207 prognostic statements were staged for future AI-assisted curation.

### Submission of Diagnostic Evidence via *ClinVar This!*

Transformed and validated data were formatted for submission to ClinVar using the GKS *ClinVar This!* tooling^19^. Under the VA-Spec, clinical significance is represented by *predicates*. Of the available evidence types (therapeutic, diagnostic, prognostic), only diagnostic statements were selected for submission. Consistent with IGM clinical workflow, these were uniformly represented using the *isDiagnosticInclusionCriterionFor* predicate, enabling direct mapping without additional predicate inference.

In total, 1,499 diagnostic statements were successfully transformed into ClinVar submission records, with 93 additional statements flagged for further clinical review. These records were validated against the ClinVar API submission schema and submitted via the GKS *ClinVar This!* interface^19^.

At the time of initial payload submission (November 2025), 1,499 fully-structured diagnostic evidence statements were deposited to ClinVar (**Figure 3a**). Among these records, there were 982 Tier I (Strong) and 518 Tier II (Potential) statements submitted. These diagnostic records covered a wide range of tumor type designations, with Embryonal Rhabdomyosarcoma and Medulloblastoma (H3 K27M-mutant) being the most frequent designations (171 and 146 statements, respectively) (**Figure 3b**).

**Figure 3.**
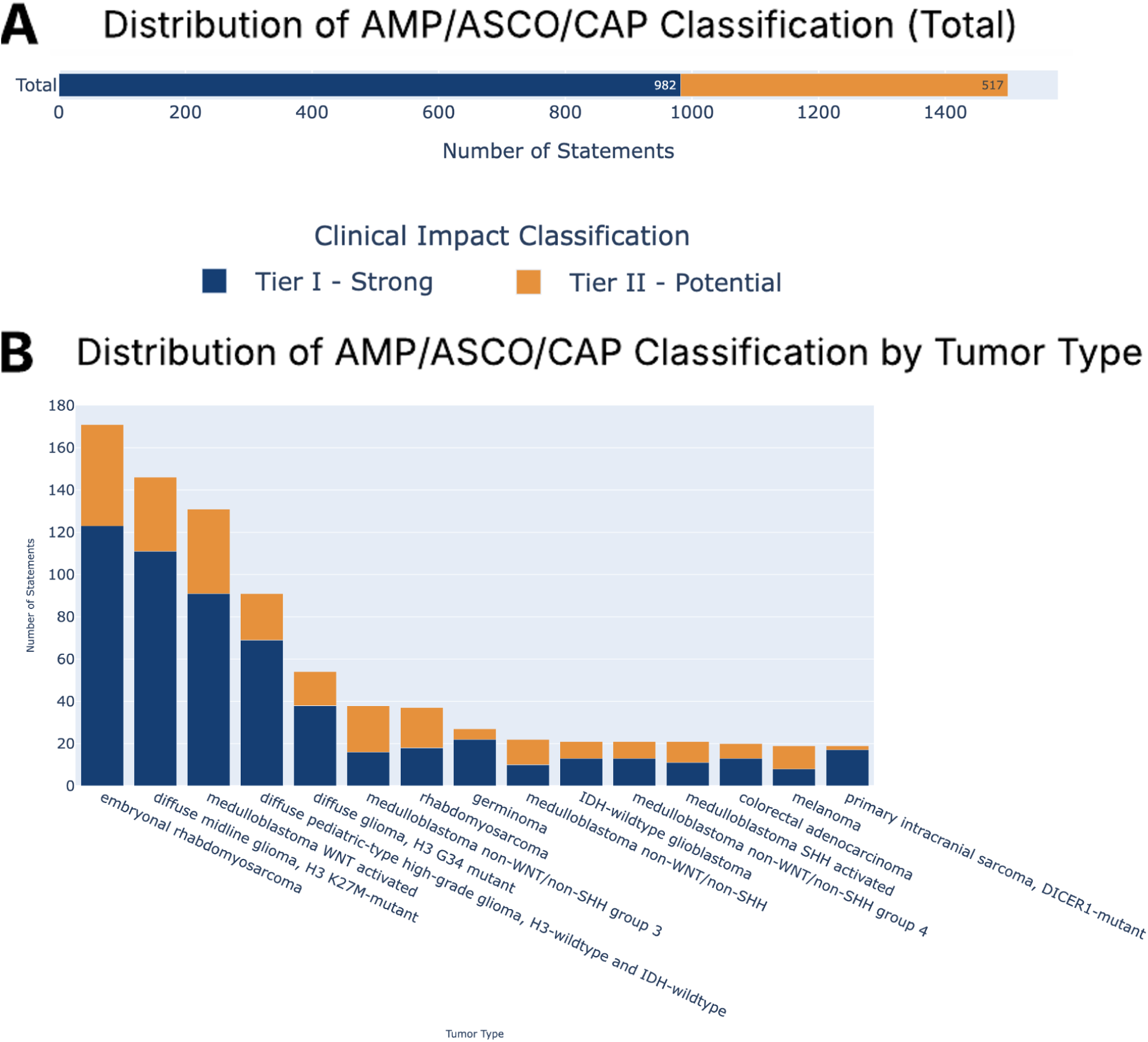
ClinVar Submission of Diagnostic Statements from Pediatric Data from Nationwide Children’s Hospital. **(A)** Distribution of 1,499 AMP/ASCO/CAP classification tiers for all statements submitted to ClinVar from the NCH data set (as of November 2025). **(B)** Tumor type coverage for evidence line submission by evidence level (only top 15 most frequent tumor types shown). Full payload available from https://github.com/GenomicMedLab/mci-knowledge-pilot.

### Classification of prognostic and therapeutic evidence lines using token dictionary matching

Clinical significance values for prognostic and therapeutic evidence lines were unable to be assigned during the parsing and transformation of variant assessment data due to the complexity of evidence representation. While the strength of evidence is indicated by AMP/ASCO/CAP Tier evaluation, the clinical significance value is essential for indicating the directionality of evidence in the VA-Spec predicate (e.g. ‘predicts worse outcome’ or ‘predicts better outcome’ for prognostic evidence). The predicate value is informed by supporting evidence that is provided by the clinical scientist during the variant assessment workflow and takes the form of free-text comments, screenshots, and identifiers for other resources (e.g. PMIDs, clinical trial IDs, etc.). While past research has demonstrated that similar classes of knowledge reporting often follow parallel grammatical patterns^20^, there may still be nuances in the variant interpretation field regarding how evidence is applied towards an outcome in two different contexts.

To evaluate automated predicate classification for prognostic and therapeutic evidence lines, we first applied naïve token-level dictionary matching to free-text comments parsed from input variant assessment files. Dictionaries were created for each predicate type for therapeutic and prognostic evidence lines: “resistance” and “sensitive” for therapeutic evidence lines, and “better outcome” and “worse outcome” for prognostic evidence lines (**Supp. Figure 4a**). Free-text comments were scanned for tokens belonging to each dictionary with predicates being assigned based on exact matches. Predicates with multiple matches and negation were also tracked. From this approach, we identified very few predicates able to be assigned for both therapeutic (156 sensitive, 0.09%, 148 resistant, 0.09%) and prognostic (568 worse, 0.32%, 120 better, 0.07%) evidence lines (**Supp. Figure 4b**). These predicate counts increase when also considering negation, however, doing so would also warrant additional, more complex methods such as sentence dependency parsing to determine where the negation is being applied. Taken together, these results underscore the need for methods that can account for context and the complexities in how variant evidence is applied.

### Development of a framework for scalable verification of applied variant evidence predicates classified by AI agents

To address the complexities and surrounding context necessary for predicate classification of evaluated variant evidence, we designed a framework to support an AI agentic curation approach. The AI agent leverages statistical pattern recognition over clinical language to generate structured, testable outputs^8,21^. As a result, our framework is designed to improve reliability and reproducibility while remaining reviewable by a human and subject to expert validation^22^.

To support the use of LLMs to perform agentic classification on evidence line data, we modeled the structure and possible enums for prognostic response evidence as distinct Pydantic classes. This generalizable framework, named Wags-LLM^22^, enables data from evaluated variant assessment forms to be assembled into a prompt, submitted, and validated for AI classification tasks as defined by evidence type. After submission of the prompt, responses from AI agents are compared against defined schemas to enable validation of data. An example of this is shown in **Figure 4**, in which evidence line descriptions and evaluator comments for prognostic evidence can be submitted for AI classification as one of four distinct, valid values: better outcome, worse outcome, unclear (e.g. prognosis mentioned but clinical significance is ambiguous), or not mentioned (e.g. prognosis is not mentioned or does not apply to the evaluated variant). Validated responses can be output as JSON for rapid downstream confirmation by domain experts and exported for internal or external use in variant assessment. The use of this framework enables the scalable, rapid assessment of clinical significance for variant allele specification data without sacrificing the expert guardrails that enable high quality, expert determined decision making for downstream patient care.

**Figure 4.**
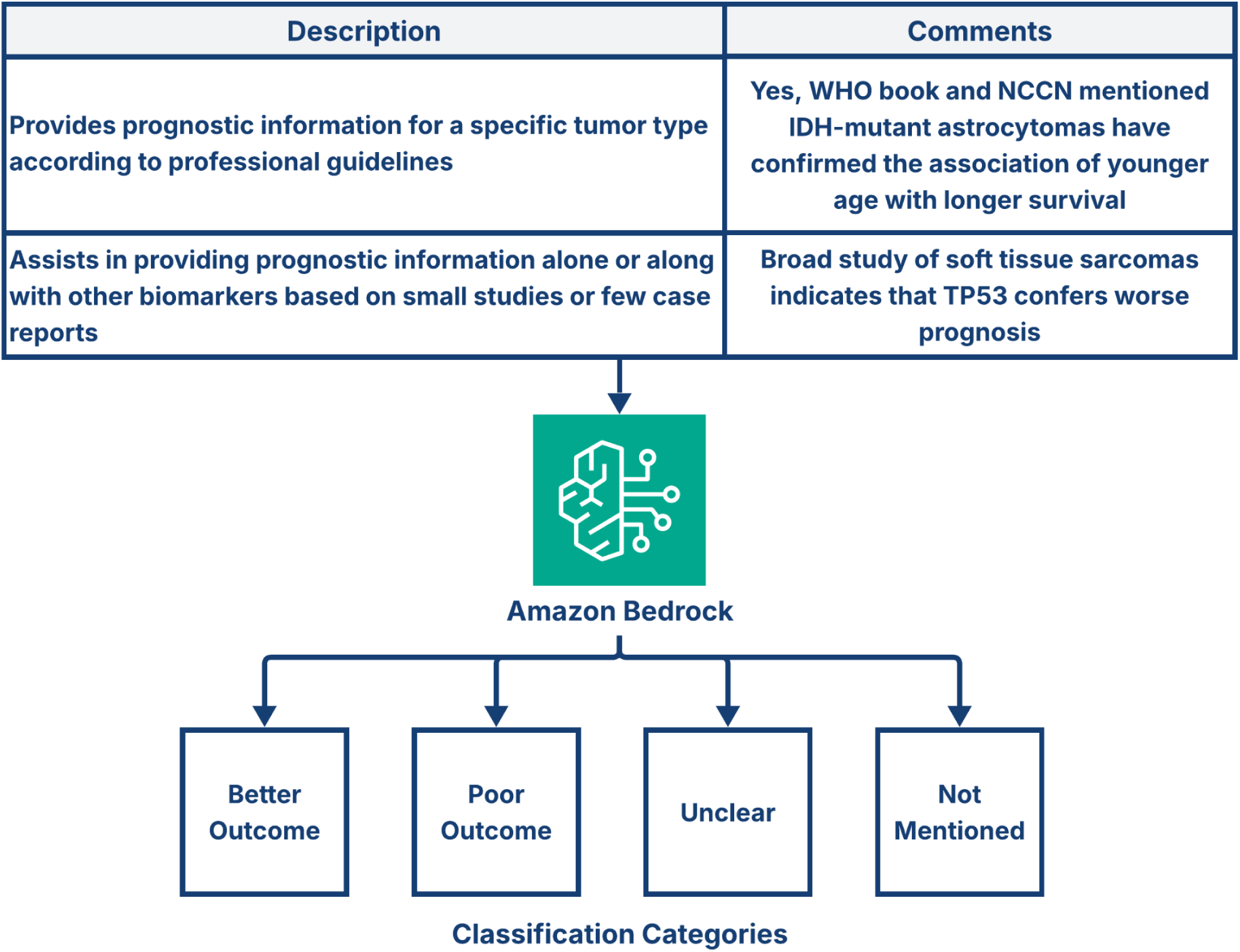
Designing a framework for AI agentic classification of VA-Spec predicates for prognostic evidence lines. Evidence line evaluation data for prognostic evidence lines from internal variant assessment forms are submitted as a classification task through Amazon Bedrock. For each evaluated evidence line, the field description and evaluator comments are submitted independently to an AI agent. The agent is tasked with classifying the clinical significance into one of four categories: better outcome, poor outcome, unclear (e.g. prognosis mentioned but clinical significance is ambiguous), or not mentioned (e.g. prognosis is not mentioned or does not apply to the evaluated variant)

### Classification of predicates for prognostic evidence lines using AI agentic curation

Each evidence item includes free-text comments that provide contextual interpretation. Prompts were assembled with free-text comments, evidence line descriptions, and prognostic class descriptions programmatically (**Figure 5**). We manually classified comments at the evidence-line level into three outcome categories: *poor outcome*, *better outcome*, or *indeterminate* (further subdivided into *null* or *unclear*). We then applied our predicate classification framework to these comments to infer clinical significance. Out of a subset of 163 evidence lines, the LLM classifications agreed with curator labels for 117 cases. Stratified by category, classification accuracy was 86.5% for poor outcome comments, 82.0% for better outcome comments, and 50.8% for indeterminate comments (**Figure 6**). Notably, the model only misclassified a poor outcome as a better outcome once, indicating strong separation between these classes. Misclassifications were more common for indeterminate comments, which were assigned to poor or better outcome categories 30 times out of 61, indicating a hallucination of sorts, proposing certain information exists in the comment that in fact does not. Although hallucinations are a well documented artifact of AI, a higher percentage of misclassification in the indeterminate comment category is consistent with the inherent ambiguity of brief, context-limited free text comments, which can be challenging even to human curators.

**Figure 5.**
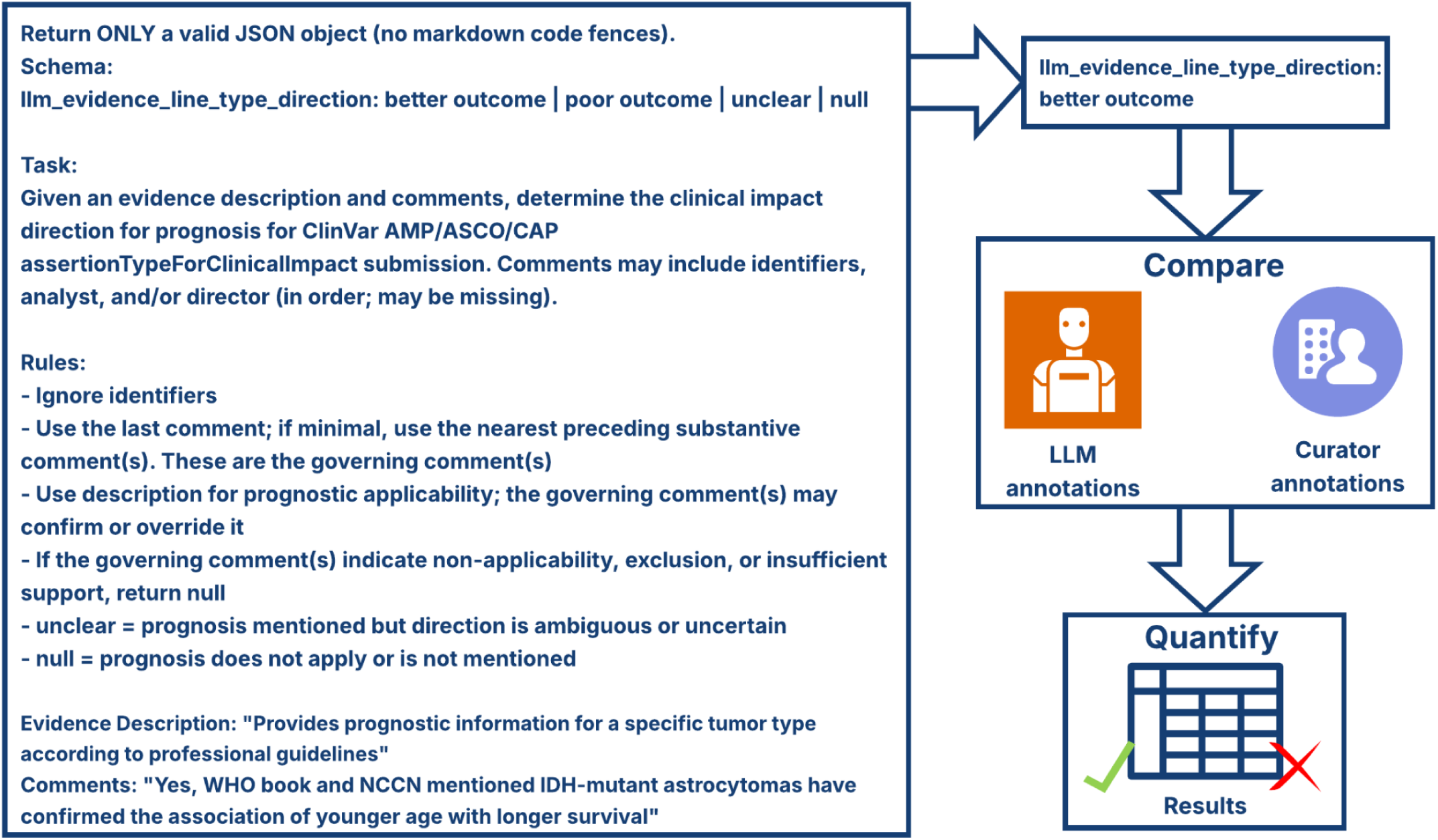
LLM prognostic direction curation workflow. Overview of the large language model (LLM)- based workflow for automated annotation of comments associated with applied prognostic evidence criteria. The structured prompt includes class definitions and outcome requirements. The annotations provided by the LLM are then compared to the curator annotations and evaluated.

**Figure 6.**
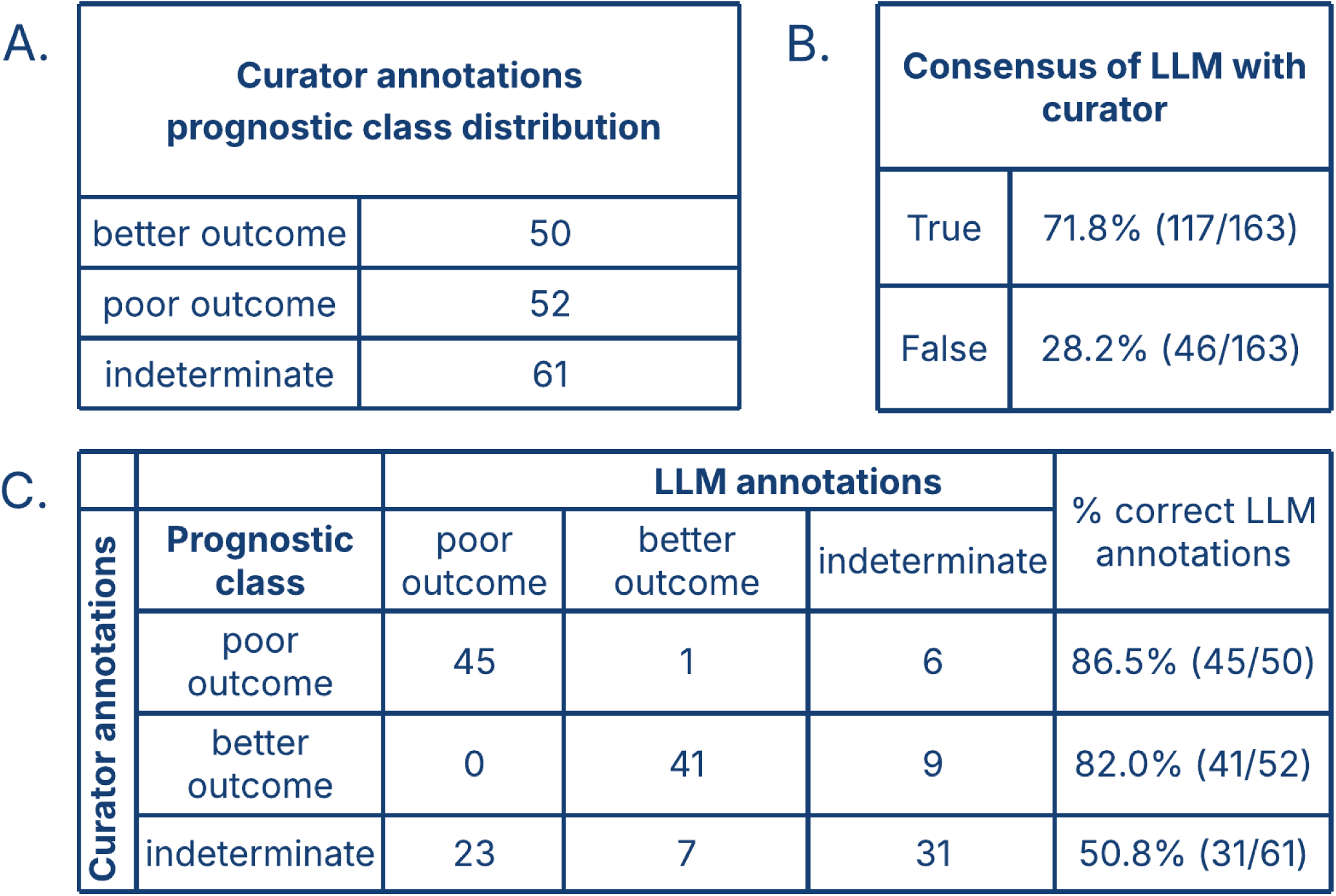
Evaluation of LLM annotations for the directionality of prognostic evidence criteria. A) Distribution of the prognostic classes in the subset of human annotated evidence criteria. B) The total number of LLM annotations in agreement with human annotations. C) A confusion matrix expanding on the LLM annotations that agreed with human annotations, by prognostic class.

## DISCUSSION

This study addresses a key limitation in clinical genomics. Although variant interpretation workflows produce large volumes of high-quality, expert curated evidence, the downstream transformation of this knowledge into standardized, computable and shareable formats remain limited. The constraint is not the curation effort itself, which is part of routine clinical evaluation of genetic testing results, but the ability to programmatically operationalize the outputs of that classification activity. While modern, structured interfaces exist for curators to provide the necessary structure to newly generated data, historical data sets stored in diverse file types require much greater manual effort. By systematically transforming these historical data sets and applying off-the-shelf agentic solutions to fill in the gaps, we demonstrate a pilot framework for streamlining transformation of data into shareable genomic knowledge.

The transformation of semi-structured variant assessment data into standardized, computable representations is a critical step towards scalable and FAIR knowledge sharing for precision oncology^23^. As shown in **Figure 1**, semi-structured variant assessment data can be reliably ingested and transformed into standardized representations suitable for downstream clinical use and sharing. By transforming variant assessment form content into structured Pydantic data models and mapping these to the GA4GH GKS specification, we pilot a practical workflow for converting locally curated variant interpretation data into validated, standardized data object that enables high-throughput dissemination of genomic knowledge.

Notably, we were able to successfully ingest 3,147 somatic variant assessment forms generated from our institution. Quality control checks revealed several consistent sources of variability and data loss related to missing or incomplete information. Forms were identified and excluded due to missing tumor type designations or lack of any applied evidence criteria. This pattern suggests that even within standardized workflows, key contextual elements may differ in structure or content (whether by unknown circumstances or special, context-specific deviations) and checks should be implemented to safeguard against this. Both of these fields are critical to both variant interpretation and GA4GH GKS representation and thus these records were deemed unsuitable for structured knowledge representation and downstream data sharing efforts without expert review and reconciliation of missing data. However, we were able to extract evidence lines from the remaining files and construct diagnostic statements as shown in **Figure 2**. Taken together, these findings highlight both the scale of available structured knowledge from routine clinical workflows and the challenges presented by incomplete data.

While structure can be applied to expected, routine fields with ease, free-text comments present a unique challenge in the field of variant interpretation. These free-text fields were found next to every evidence line as an optional annotation that may or may not inform the clinical significance value or interpretation of the applied evidence line. Of these three extracted evidence line types, we found diagnostic evidence line interpretation to be fairly straightforward where GA4GH GKS VA-Spec predicates could be assigned solely with structured data, independent of any free-text comment-derived context. Thus, in our workflow we assigned these predicates, transformed them into ClinVar submission format, and submitted them for public data sharing (as demonstrated by our November 2025 submission in **Figure 3**). This ease of assignment was not the case for therapeutic and prognostic evidence lines, as we found free-text comments could contain additional information critical to the clinical significance of application (e.g. whether a prognostic evidence line indicates a better or worse outcome for an individual). In these cases, we determined that additional methods would be needed in order to accurately structure the data.

In order to add structure to data and correctly assign GA4GH GKS VA-Spec predicates to variant evidence, we utilized AI agentic curation efforts. As demonstrated in **Supp. Figure 4**, assigning predicates on free-text comments is too complex a task to be accomplished through simple methods, such as dictionary look-ups which only recover a small fraction of predicates due to the variability and contextual dependence of clinical language. In contrast, structured data curation by AI agents accelerates the inference of missing data from free-text comments and thus operates as a scalable approach towards curating variant clinical significance. Accordingly, we designed a framework to support AI agentic classification of prognostic evidence in a modeled, validatable format. Individual evidence lines and their free-text comments could be assessed by an off-the-shelf AI agent with responses able to be validated as one of four possible responses. Under this schema, individual evaluated evidence lines from a single variant assessment can be assessed and “rolled up” to form a predicate classification for the entire statement. In our evaluation of 163 randomly sampled prognostic evidence lines with free-text comments, we demonstrated that the AI curation agent was reliable for predicting ‘poor outcome’ and ‘better outcome’ VA-Spec predicate classifications with 86.5% and 82.0% classification accuracy respectively. We observed a strong separation between these two classes while having trouble with ‘indeterminate’ classifications. We found in this analysis that, despite the exact nature of the wording, as long as the outcome is well-described in a human readable manner, the correct outcome would be assigned. Comment descriptions that were too vague or made references to other cells (e.g. comments such as ‘see above’ or ‘described previously’) were frequently unable to be accurately determined. Similarly, comments which made reference to attached screenshots or external evaluations remained difficult to accurately predict a predicate. Our curator consensus rate agreed with the AI 71.8% of the time, further suggesting confidence in LLM classifications of ‘better outcome’ and ‘worse outcome’. With the greatest source of disagreement coming from ‘indeterminate’ records, our results suggest that a flagging or consensus mechanism could be put into place for indeterminate records to enable efficient direction of human effort. By quickly binning explicit ‘poor outcome’ and ‘better outcome’ records, valuable human curator time can be used instead to resolve indeterminate cases.

While our results are encouraging, the use of large language models introduces concerns regarding data privacy for protected health information (PHI). Although our variant interpretation dataset was curated to exclude direct identifiers, there is still the risk of identifying information being embedded within free-text fields. These fields may contain undetected direct identifiers, or more likely, contain rare or uniquely identifying details (e.g. rare cancer subsets or characteristics) that may inadvertently be retained and memorized by the model if the proper safeguards aren’t in place^24,25^. These risks are especially prescient when using external or commercial models, where data retention and model training practices introduce further risk of unintended PHI exposure. To mitigate these concerns, we would suggest implementations of this framework to use de-identified datasets within institutionally controlled environments with strict data governance and zero-retention policies. These considerations emphasize that while AI agentic curation can provide scalability, its application must be carefully constrained to ensure patient privacy and compliance with PHI protection regulations.

This work demonstrates a pilot framework for transforming semi-structured variant assessment data into computable clinical assertions using GA4GH standards and AI-assisted curation. Our results highlight that while variant assessment data commonly has structured fields that can be readily mapped, there are complexities to the data that can complicate scalable knowledge representation. We show that this limitation can be alleviated through validatable AI agentic approaches, freeing up valuable human curation time for where it is most needed. While challenges still remain (e.g. interpreting vague or incomplete annotations), our pilot approach provides a path forward for integrating AI into clinical variant curation/deposition workflows for data sharing at scale. By improving the handling of indeterminate cases and expanding to additional evidence types, our framework represents a step towards improving the scalability and availability of genomic knowledge.

## MATERIALS AND METHODS

### Generation of Variant Assessment Forms

Variant assessment forms are structured documents used to capture the evidence, criteria application, and final classification of genomic variants in a standardized and auditable format. Within the clinical workflow, variant assessment forms function as the primary record of variant classification, encoding the evaluation of genomic, computational, and literature-derived evidence in accordance with established clinical guidelines. For germline variants, assessments are performed following ACMG/AMP standards^26^, while somatic variants are evaluated using AMP/ASCO/CAP Tiering frameworks and, where applicable, more recent ClinGen/CGC/VICC oncogenicity criteria^27^. Historically, evidence and criteria were recorded in Excel-based variant assessment templates, ensuring consistent application of classification rules and traceability of interpretive decisions.

Clinical variant scientists generate variant assessment forms as part of the primary variant analysis workflow following sequencing and initial filtering. Candidate variants are identified through systematic review of annotated result sets, prioritized using population frequency, variant allele frequency, clinical databases, and disease-relevant gene lists, and visually inspected in Integrated Genomics Viewer (IGV) to exclude technical artifacts. For variants selected for formal interpretation, the variant scientists compiled multi-source evidence, including population databases, disease-specific repositories, functional data, and literature, and document these data within the variant assessment form by applying predefined evidence criteria and classification rules. The completed form includes structured evidence annotation, classification rationale, and supporting documentation such as database queries and visualization screenshots, and is stored in a standardized file system for downstream use.

Following completion, variant assessment forms undergo clinical director review as part of the clinical sign-out process. Clinical directors evaluate the underlying evidence, verify correct application of classification criteria, and confirm consistency between the variant assessment form and the reported variant interpretation. Where necessary, revisions are made and documented prior to approval. Director approved forms are uploaded to the clinical workflow system, and linked to the individual report, with only director-approved variants eligible for reporting. This review and sign-out process ensures that variant classifications are accurate, reproducible, and compliant with clinical laboratory standards before communication to ordering providers.

### Data Availability

GKS-formatted submission files are publically accessible from: https://github.com/GenomicMedLab/mci-knowledge-pilot.

### Ingest of Variant Assessment Forms

Variant assessment forms were programmatically accessed and processed using a three-step workflow (**Supplementary Figure 2**). Completed variant assessment forms were extracted into a Polars DataFrame. Ingested data was cleaned and normalized to disease concepts (see GKS mapping and disease normalization). Normalized, cleaned data was validated and represented using Pydantic models for variant assessment data objects.

### GKS mapping of ingested Variant Assessment Forms to GKS VA-Spec AMP/ASCO/CAP 2017 profiles

Ingested variant assessment data objects were then transformed to GA4GH GKS Pydantic models using the Python reference implementations (VA-Spec Python for representing evidence and VRS-Python for representing variants). Variants were translated into their corresponding VRS Alleles (**Supp. Figure 3**). Evidence lines where requirements were adequately met were collated together and used to generate Variant Annotation Specification (VA-Spec) AMP/ASCO/CAP 2017 compliant Variant Clinical Significance statements. This process was done for each sheet (most recent assessment) for diagnostic evidence. Resulting outputs were categorized according to the completeness of GKS transformation under the following enums: Success, Flagged for Clinical Review, Director Not Approved, Failed.

### Submission of GKS mapping variants using *ClinVar This!*

Outputs that were successfully transformed to GKS representations were prepared for submission to ClinVar. GKS representations were mapped to the ClinVar Submission API schema. After validation against the schema, records were submitted using the ClinVar Submission API. The code and schema for GKS *ClinVar This!* is available from: https://github.com/clingen-data-model/clinvar-this.

### Normalization of Diseases

Diseases referenced during the ingest of variant assessment forms were subject to normalization during the ingest process. Terms were programmatically compared to ontologically established concepts. Diseases without suitable concepts were mapped using a curated internal disease mapping. Similarly, if a tumor type field was previously curated by an internal clinical expert, that curated value was prioritized for use in the disease normalizer. The code for normalization services are available from:

Disease: https://github.com/cancervariants/disease-normalization.

### Implementation of VRS Models using VRS-Python

Variant representation specification compliant models were implemented using the VRS-Python language support toolkit and reference implementation^28^. The VRS-Python toolkit is available from: https://github.com/ga4gh/vrs-python/

### Notebook Availability

All quantification and analysis work was performed using Jupyter Notebooks. Data and analyses are available for public access via: https://github.com/GenomicMedLab/mci-knowledge-pilot. Additionally, since the November batch submission, we have updated our GKS representations to require a VRS Allele, not just the corresponding HGVS expression. Therefore, there may be discrepancies in newer versions (post-November 2025) of the GKS-formatted data.

### VA-Spec predicate classification of evidence lines using token-level dictionary matching

Token dictionaries were created for expected VA-spec predicate types for therapeutic and prognostic evidence lines. A token dictionary was also created for common negation words. Free-text comments from each GKS transformed variant assessment form were scanned for tokens belonging to their corresponding evidence type (e.g. therapeutic evidence was compared against therapeutic token dictionary). Word matches and instances of negation were quantified and used to naively assign predicates. Objects with multiple opposing word matches were also quantified (e.g. both ‘sensitive’ and ‘resistant’ appear in the same comment).

### Implementation of Framework for AI agentic curation

Wags-LLM is a lightweight framework for orchestrating large language model workflows, providing support for versioned prompts, Pydantic-based output validation, and optional caching^22^. It is designed to improve reliability and reproducibility while remaining easily extensible through user-defined prompts and response schemas. Wags-LLM was utilized for structuring prompts, submission, and validation of downstream AI agentic curation tasks.

### VA-Spec predicate classification of evidence lines using applied AI agentic curation

Each evidence item may include associated free-text comments contributed by analyst(s), directors, or both. Comments encompass literature citations, questions, clarifications, summaries, and approvals. In the source Excel document, comments are linked to an evidence line if they appeared in the same row, with each comment stored in a separate cell. For analysis, comments associated with a given evidence line were aggregated into a list.

A subset of forms to test the LLM was randomly sampled until at least 50 manually annotated comment lists were obtained for the better and poor prognostic classes. Manual annotations were restricted to comments associated with applied prognostic evidence lines.

To classify clinical significance, we constructed a templated prompt that included: (i) a defined professional role for the model, (ii) explicit descriptions of each of the prognostic classes, (iii) a required outcome schema, and (iv) placeholders for the evidence line description and associated comment list. Prompt variants were iteratively refined and versioned numerically to support systematic evaluation and review.

LLM-based classification was performed using the Claude Sonnet 4.6 model at a temperature of 0.0 for AI curation of prognostic classes. Result stability was verified by analyzing model performance across three independent experiments using identical samples and prompt. Model performance was evaluated by agreement with curator-assigned labels and summarized using a confusion matrix. For analysis, the “unclear” and “null” classes were consolidated into a single “indeterminate” class.

---

**Supplementary Figure 1.**
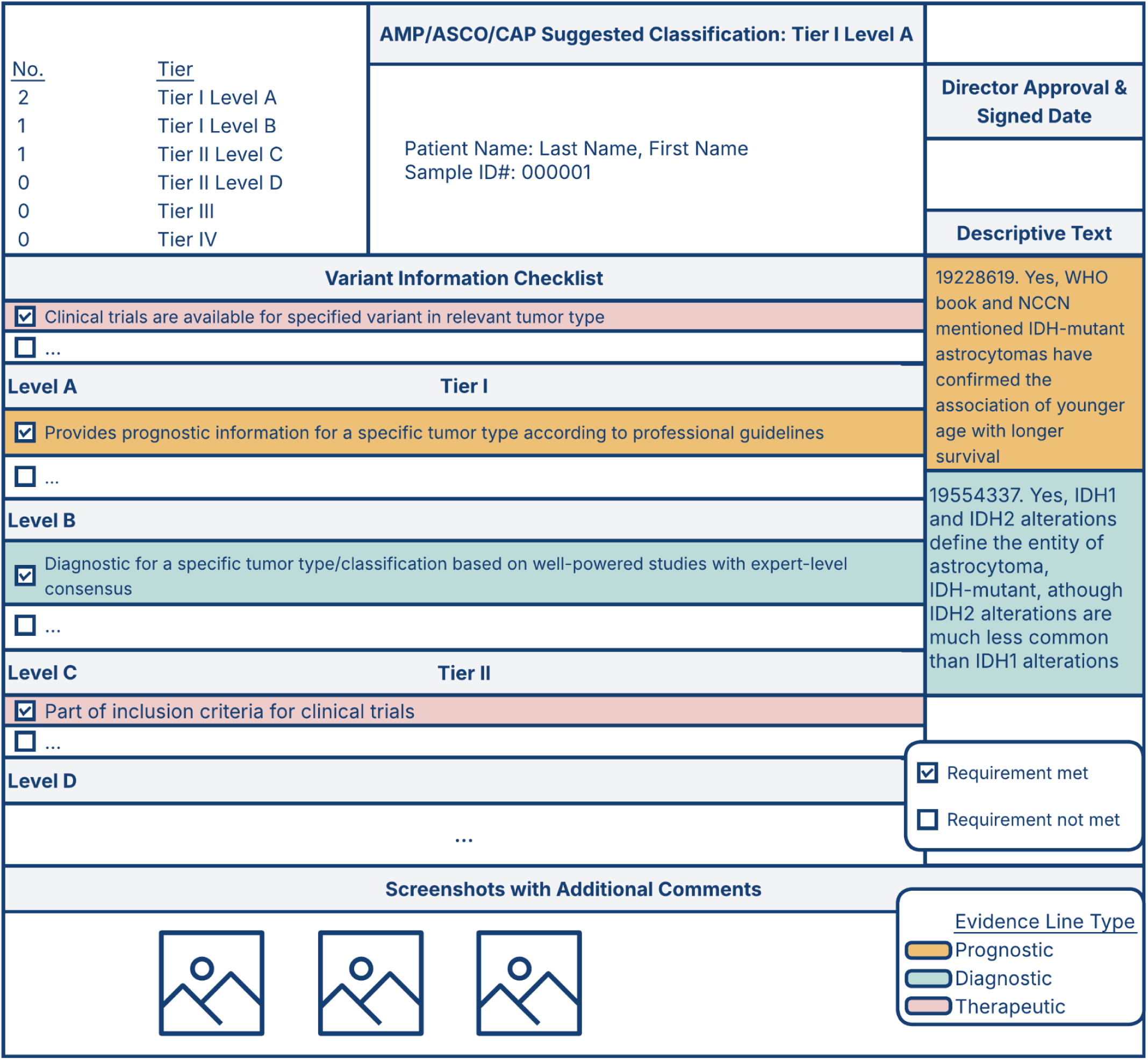
Example of Somatic Variant Assessment Form. Variant assessment form used by variant analysts, scientists and clinical directors for the classification of cancer-associated variation under the AMP/ASCO/CAP 2017 classification SOP. Variant evidence can be applied as therapeutic, diagnostic, and prognostic evidence lines.

**Supplementary Figure 2.**
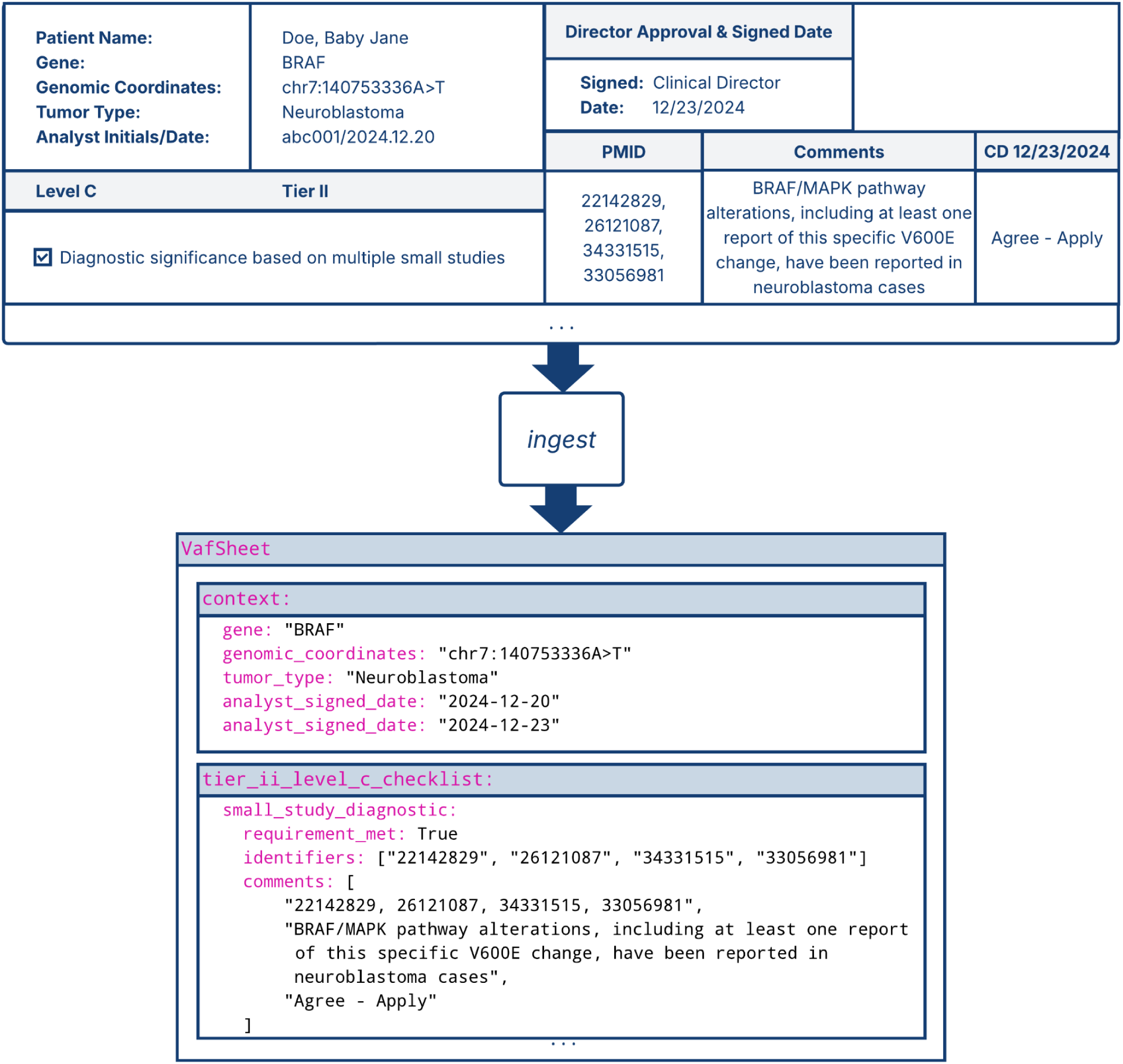
Ingestion of Variant Assessment Forms into Pydantic Model. Historical variant assessment forms stored as excel documents are ingested and transformed into Pydantic objects.

**Supplementary Figure 3.**
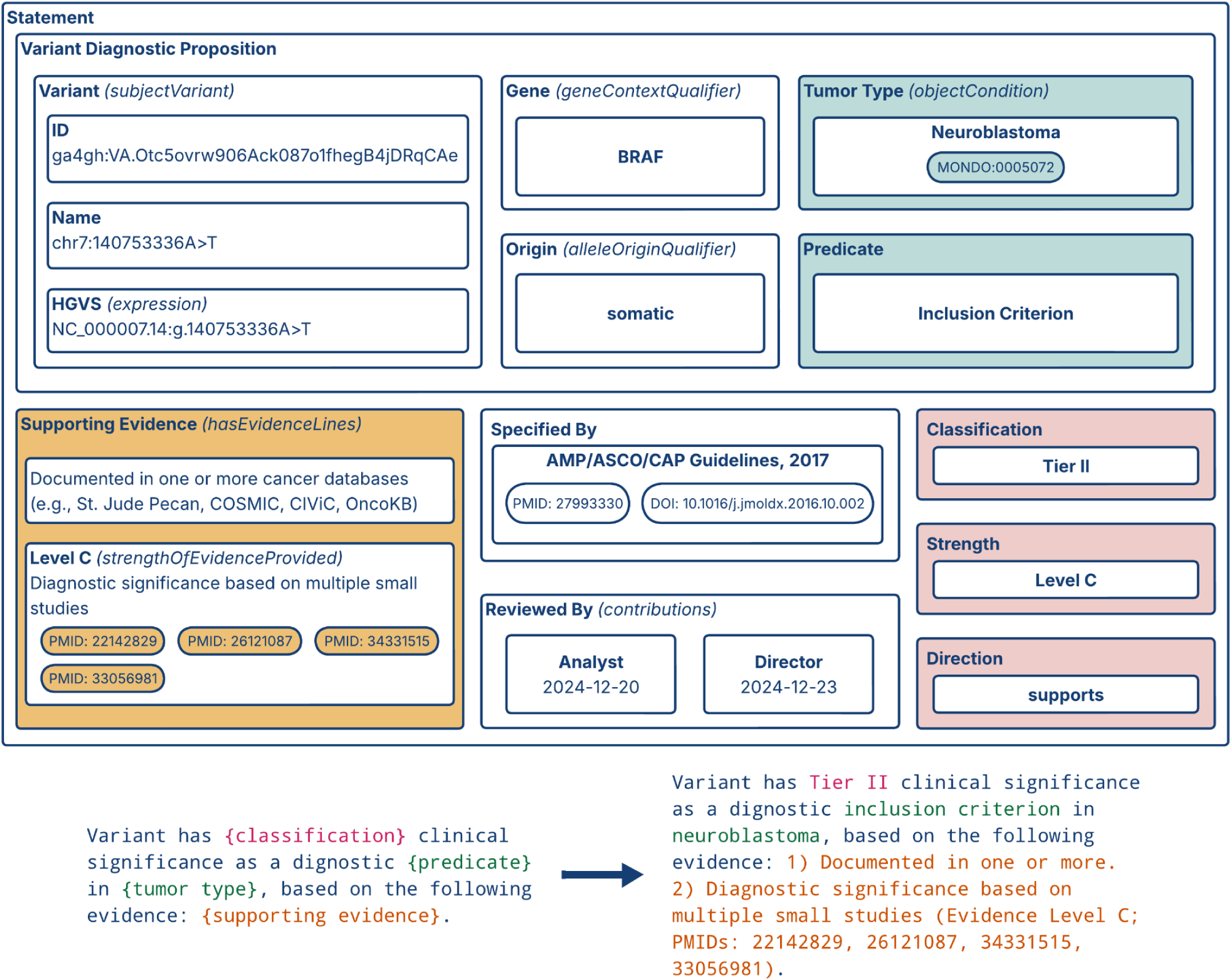
Transformation of Variant Assessment Form derived data into Variant Annotation Specification evidence statements. Data is further transformed into VA-Spec validated evidence statements for downstream knowledge sharing.

**Supplemental Figure 4.**
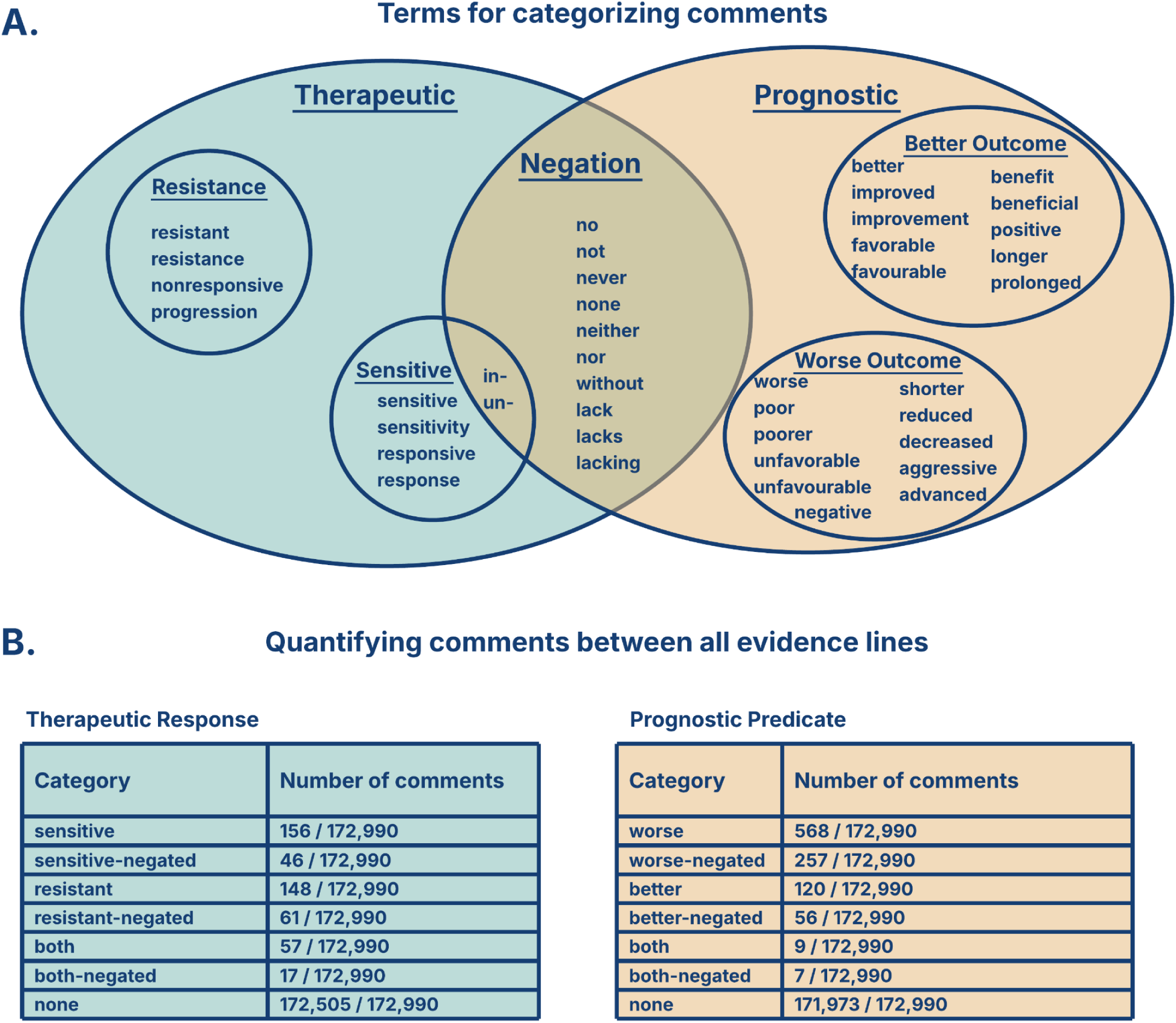
F**r**ee **text comments cannot be categorized based on key vocabulary terms. A)** Therapeutic comments were divided into those attributing resistance or sensitivity as effects of the variant. Lack of sensitivity does not result in resistance and therefore the “in-” and “un-” prefixes are sensitive-negated. **B)** Prognostic comments were divided into better and worse outcomes. Negation of the word “favorable” directly correlates with worse outcomes, not just “better-negated” outcomes and therefore is in the worse outcome group. There were no comments that had conflicting terms within the therapeutic and prognostic categories. There can be multiple comments for one evidence line. Therapeutic and prognostic analyses were completed independently, a comment may contain both therapeutic and prognostic content.

**Supplementary Tables - Transforming Semi-structured Variant Assessments int…**

**Supplementary Table 1. Field descriptions for Pydantic model of pediatric variant assessment data.**

